# ‘The Association of Scoliosis Properties with Spinal Cord Tethering: A Statistical Model for Prognostication’

**DOI:** 10.1101/2022.03.10.22272232

**Authors:** Kourosh Karimi Yarandi, Esmaeil Mohammadi, Abbas Amirjamshidi, Mohammad Shirani Bidabadi, Ahmad Pour Rashidi, Sina Azadnajafabad, Seyed Farzad Maroufi, Maisam Alimohammadi

## Abstract

**Objective:** To evaluate the relationship between the structural measures of scoliosis and underlying spinal cord tethering (SCT) and proposing a statistical prognostication model.

**Study design:** Cross-sectional.

**Setting:** Academic healthcare center

**Methods:** 128 definite scoliosis cases that were candidates for corrective surgery were enrolled. Anterior-posterior whole column digital radiographs and whole-spine MRI (supine for all samples and adjuvant prone MRI for suspected cases with tight filum terminal) were performed. Univariate and multiple logistic regression were used for the analysis of association and interaction. Association of SCT with structural features of scoliosis –Cobb angle, convexity, and type (idiopathic and congenital)– age, and sex were assessed.

**Results:** None of the study variables showed a statistical association with SCT in univariable and multiple logistic regressions. After inclusion of Cobb angle-convexity-type interaction, higher Cobb angle, idiopathic scoliosis, dextrosoliosis, and male gender had a significant effect. Stratification for convexity discovered a positive association of Cobb angle and SCT in idiopathic patients with dextroscoliosis (1.02 [1.01–1.03], 0.049). In contrast, in congenital cases, the rate of SCT decreased by higher left-sided Cobb angles but it was not statistically significant (0.94 [0.88–1.01], 0.104).

**Conclusion:** The risk of spinal cord tethering was not zero in any of the subgroups and no SCT-free group could be detected. Conventional MRI should be preoperatively performed for every case of scoliosis and thoroughly examined for signs of tethering. Clear imaging of patients at higher risk of SCT should not be decisive and further workup should be utilized before proceeding with reconstructive surgery.

## Introduction

Scoliosis is defined as a three-dimensional spinal deformity involving one or more spinal curves with lateral deviation and axial rotation of the vertebrae.(1) This condition is usually classified into two major etiologic categories: idiopathic and congenital.(2) Congenital scoliosis (CSc) arises from a type of vertebral malformation, e.g. hemivertebra(3), while in the idiopathic form (ISc) no other underlying anatomical problem can be identified.(4, 5) Other disorders can occur along with both CSc and ISc and they are more frequently seen in CSc.(6-10) Tethered cord syndrome is highly correlated with scoliosis.(11-15) For those patients with symptomatic or subclinical cord-tethering problems, a cord-releasing procedure before realigning surgery is proposed.(16) During realignment, tension can be imposed on the spinal cord, potentially causing a neurological deterioration. As a result, recognition of such conditions should be carried out before reconstructive surgery.(17-19)

Based on the side of the greater convexity of scoliosis, it can be further subdivided into levoscoliosis and dextroscoliosis. If the convexity of the greater curve of deformity is faced to the right, the condition is known as dextroscoliosis and if it is to the left, the case will be defined as levoscoliosis. Previous studies claimed that dextroscoliosis can be discovered in 60% of all scoliotic spinal columns.(20) Subjects with levoscoliosis have been shown to follow a more progressive course and worse surgical outcomes.(21-23) It is claimed that levoscoliosis is more frequently associated with other diseases and conditions.(21-23) In many centers a routine pre-operative magnetic resonance imaging (MRI) is used to investigate the presence of potential underlying spinal cord problems. However, a growing number of authorities have challenged the necessity of performing MRI in every scoliosis case as a routine pre-operative diagnostic tool. (24, 25) In addition, even MRI could be suspicious in some of the cases and detection of spinal cord tethering might not be straightforward, requiring additional tools for precise diagnosis. These complementary measures include urodynamic study (UDS), lumbosacral MRI in prone position, etc.(18) Here we tried to describe a method to identify more susceptible subgroups, in whom thorough examination is more justified based on their scoliosis characteristics.

## Material and Methods

### Data collection

All the scoliotic patients, who were referred to Sina tertiary Hospital, affiliated with Tehran University of Medical Sciences (TUMS), have been actively incorporated into our registry system and spine deformity database. Consecutive cases were selected according to our inclusion and exclusion criteria from this database starting from May 2013 until December 2018. Afterward, informed written consent was obtained from all of the eligible cases. All the ISc and CSc cases who were candidates for surgical correction of the deformity and having a Cobb angle of at least 10 degrees were included. Exclusion criteria were: any surgery on the spine in the past, paralytic, degenerative or other types of scoliosis, presence of double major curves, history of myelomeningocele or other open dysraphisms, any issues about the reliability of examination or interview such as mental retardation or lack of compliance, age under 3 years to exclude the infantile types, any contraindication for performing MRI such as cardiac pacemakers, any reason to preclude performing standing whole spine radiographs, and pregnancy. Demographic data and information of clinical examination were extracted. This study has been approved by the Institutional Review Board of our center.

### Paraclinical evaluation

The entire included subjects underwent a supine whole-spine MRI in addition to a standing anteroposterior, lateral, and bending whole spine radiograph. All of the imaging procedures were performed in our hospital’s radiology department. Spine radiographs were used for manual measurement of the Cobb angle (as an indicator of scoliosis severity).(26) Convexity of scoliosis was defined as the side of the greater curve of scoliosis (dextroscoliosis vs levoscoliosis). MR imaging was performed using a 1.5 Tesla Siemens equipment and analyzed by an expert team consisting of a neurosurgeon and a radiologist. Anomalies such as low lying conus medullaris (CM) at levels below L2/L3 disk space, thick fatty filum terminale (FT; diameter more than 2 mm), any type of split cord malformation (SCM, type I- and II), cord neoplasms, dermal sinus tracts, and syrinx were examined. The presence of any vertebral malformations was also analyzed for evaluation of scoliosis type (congenital vs. idiopathic). Any condition that potentially adheres the spinal cord to adjacent structures or any sign of low-lying CM was defined as SCT including the tip of CM at any level below L2/L3 disk space, presence of the neurologic signs of cord tethering [e.g., back pain, lower limb pain, claudication, paresthesia, paresis, sphincteric problems, disorganized gait, etc.] even when the end of CM is higher than L2/L3 level, thick fatty FT, SCM type I, tethering cord neoplasms, or dermal sinus tracts. A posterior locating FT in sagittal lumbosacral MRI suggests the risk of tightness. Whenever the tightness of the FT was suspected, a complimentary MRI was performed in the prone position and if the FT had a movement less than 10 percent of the total canal width in such maneuver it was also regarded as SCT.(27) It should be emphasized that we included the presence of “clinical” or “radiologic” manifestations of spinal cord tethering in this definition while in routine practice the rate of SCT is often found to be lower.

### Statistical analysis

Analyses were conducted using SPSS [IBM Corp. Released 2013] and R statistical program v3.6.0 [R Core Team (2019)]. Categorical variables such as scoliosis type, convexity, and SCT were reported by their absolute (numbers) and relative (percent) values. Continuous variables, like Cobb angle and age, were described with mean and standard deviation (SD). SCT’s association with other categorical variables was examined using binary logistic regression. The distribution of SCT subtypes in levo- and dextroscoliosis (8 × 2 contingency crosstab) was examined with Monte-Carlo’s simulation. Multiple logistic regression analysis was utilized to assess the association of SCT with sex, age, convexity, scoliosis type, Cobb angle, and three-way interactions of Cobb angle - convexity - scoliosis type.

To categorize Cobb angle (as a continuous variable) into high/low dichotomy, a productive function (as of sensitivity × specificity) was implemented on Receiver Operator Characteristic (ROC) coordinates to search for the highest yielding number and its respective Cobb angle for interactive subcategories.(28, 29) We reached to 60.5 degrees for idiopathic dextroscoliosis and 46.5 for congenital levoscoliosis samples. The latter Cobb angles were defined as the best-discriminating cut-off points. This analysis was only performed for ROC curves with a statistically significant area under the curve (AUC). Based on the interaction analyses, we divided participants into subcategories (i.e., high/low angle × levo/dextro × idiopathic/congenital scoliosis subtypes). The significance level for statistical analyses was set at P-value < 0.05. Odds ratios (OR) with confidence interval (CI) were calculated with 95% certainty.

## Results

A total number of 189 cases with scoliosis were initially evaluated and 128 patients, who were eligible according to inclusion and exclusion criteria were enrolled. Among them, 41 (32%) were male and 87 (68%) were female (M:F ratio of 1:2.1; Table 1). The mean age of patients at the time of the first visit was 15.7 ± 7.1 years. The mean Cobb angle at the time of first evaluation was 63.5 ± 27.5 degrees with a range of 10-162 degrees.

**Table 1.**
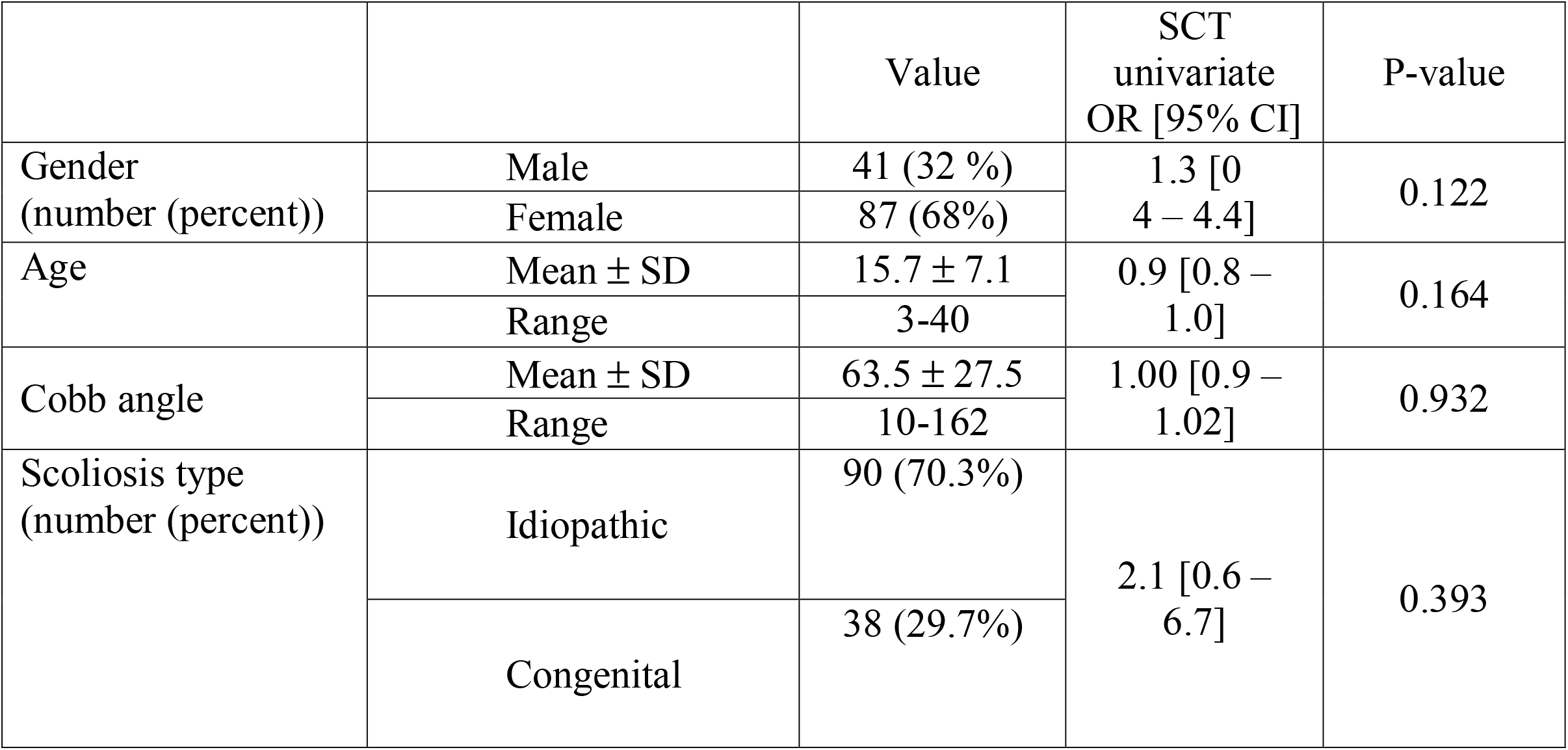
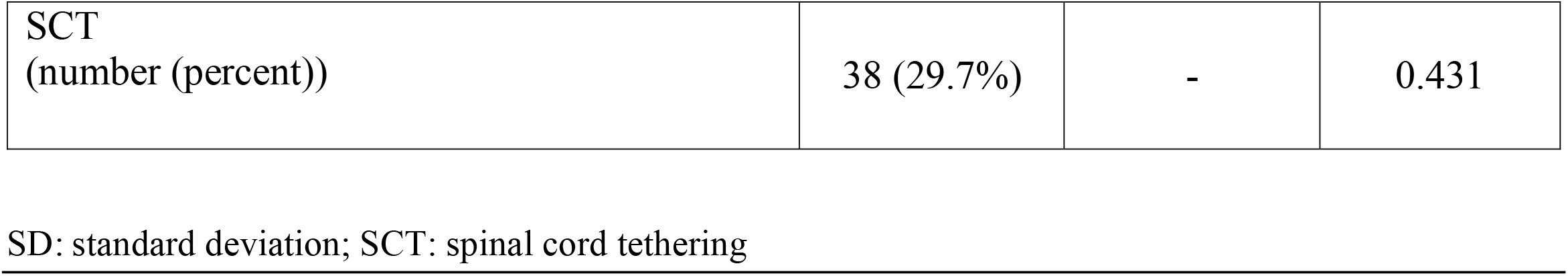
Demographic and characteristics of participants

Binary logistic regression and multiple logistic regression revealed that there is no significant relation between SCT and age, gender, Cobb angle, or scoliosis type, and convexity of scoliosis (Table 2). On the other hand, after inclusion of the interaction of Cobb angle - convexity – scoliosis type, this new variable demonstrated a significant effect of higher Cobb angle (odds ratio 1.04 [95% confidence interval 1.01–1.08], P-value=0.011), idiopathic scoliosis (0.08 [0.13 – 0.5], 0.009), dextrosoliosis (9.8 [1.7–56.8], 0.010), and male gender (2.6 [1.001–6.7], 0.05). To better delineate this effect, we divided samples into two main subgroups based on convexity. Tethering pathologies distribution did not differ between levo- and dextroscoliosis (Table 3; Monte-Carlo’s P-value [95% CI] = 0.309 [0.300 – 0.318]). Subgroup analysis discovered a positive association of Cobb angle and SCT in dextroscoliosis (1.02 [1.01 - 1.03], P-value = 0.049, Figure 1), countered by a negative association in levoscoliosis subgroup (0.94 [0.88 – 1.01], P-value = 0.104) although the latter did not reach to the significance threshold. This indicates that more severe scoliosis cases are associated with a higher rate of SCT in idiopathic dextroscoliosis patients and lower rates in congenital levoscoliotics.

**Table 2.**
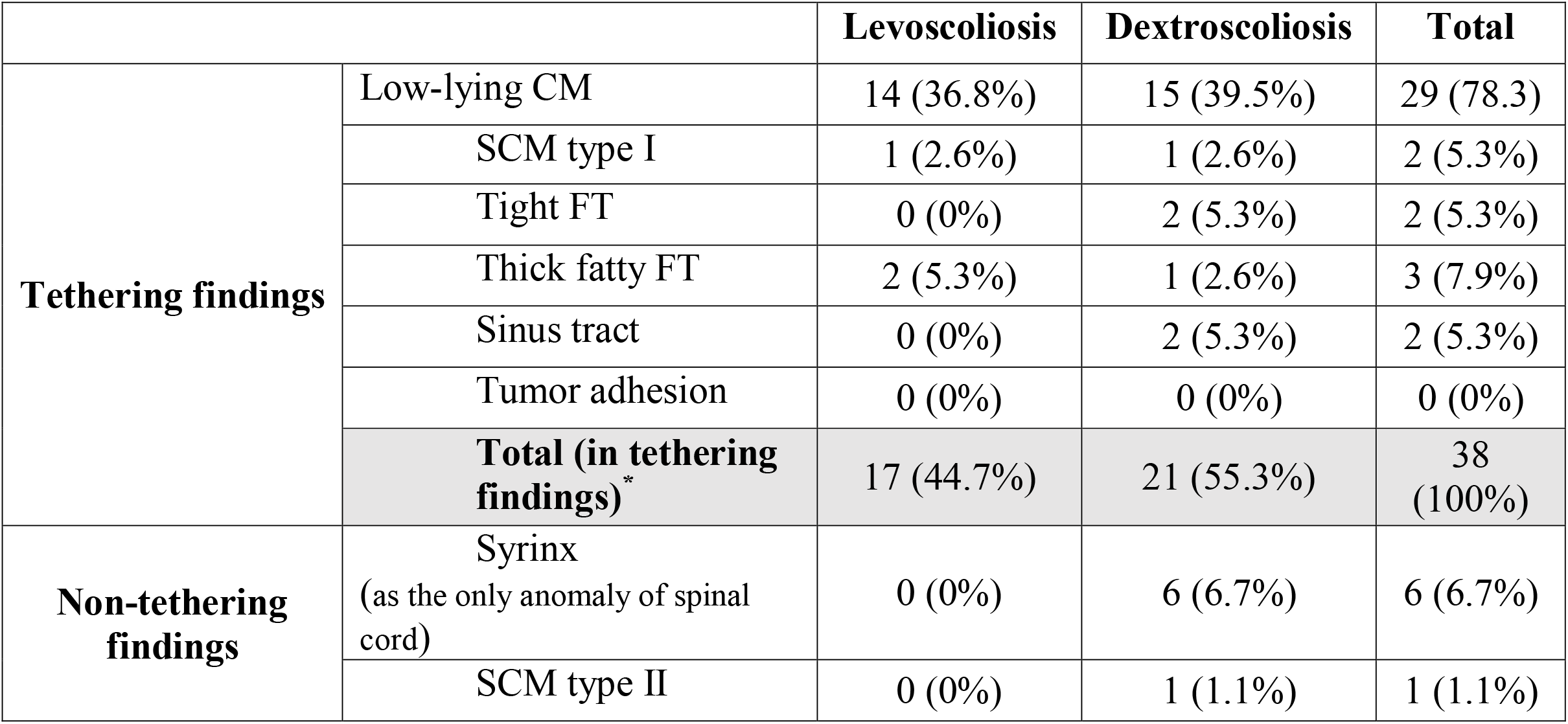
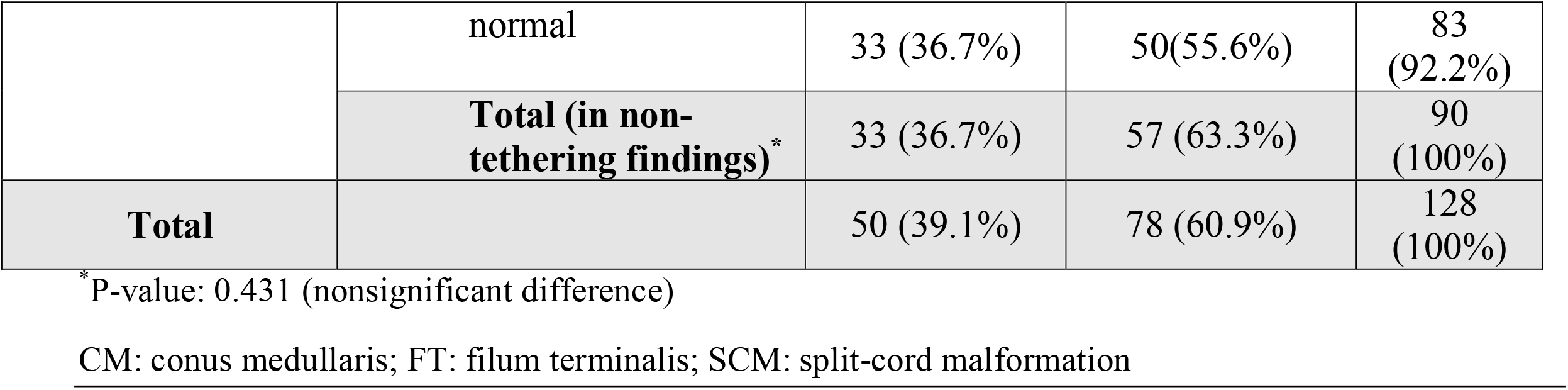
Detailed types of cord tethering etiologies in levo- and dextroscoliosis

|  |  | Levoscoliosis | Dextroscoliosis | Total |
| --- | --- | --- | --- | --- |
| **Tethering findings** | Low-lying CM | 14 (36.8%) | 15 (39.5%) | 29 (78.3) |
|  | SCM type I | 1 (2.6%) | 1 (2.6%) | 2 (5.3%) |
|  | Tight FT | 0 (0%) | 2 (5.3%) | 2 (5.3%) |
|  | Thick fatty FT | 2 (5.3%) | 1 (2.6%) | 3 (7.9%) |
|  | Sinus tract | 0 (0%) | 2 (5.3%) | 2 (5.3%) |
|  | Tumor adhesion | 0 (0%) | 0 (0%) | 0 (0%) |
|  | **Total (in tethering findings)\*** | 17 (44.7%) | 21 (55.3%) | 38 (100%) |
| **Non-tethering findings** | Syrinx<br>(as the only anomaly of spinal cord) | 0 (0%) | 6 (6.7%) | 6 (6.7%) |
|  | SCM type II | 0 (0%) | 1 (1.1%) | 1 (1.1%) |
|  | normal | 33 (36.7%) | 50(55.6%) | 83<br>(92.2%) |
|  | **Total (in non-tethering findings)\*** | 33 (36.7%) | 57 (63.3%) | 90<br>(100%) |
| **Total** |  | 50 (39.1%) | 78 (60.9%) | 128<br>(100%) |

\*P-value: 0.431 (nonsignificant difference)
CM: conus medullaris; FT: filum terminalis; SCM: split-cord malformation

**Figure 1:**
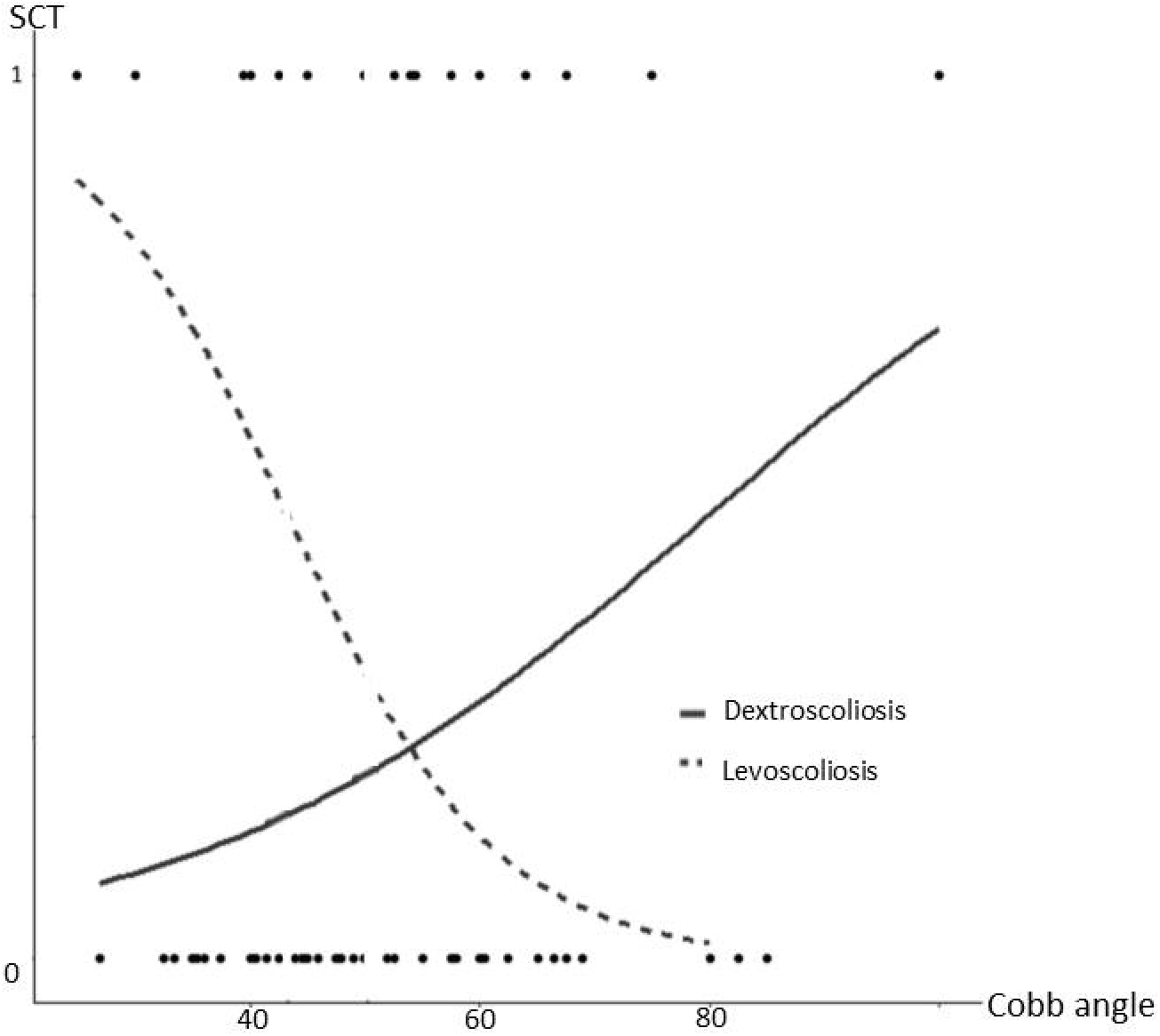
The logistic regression fit line of spinal cord tethering (SCT) and Cobb angle in two main groups of study (levo- and dextroscoliosis)

ROC curves showed different patterns in interactive subcategories and only in one of six (interactive combination of idiopathic dextroscoliosis) reached to statistical significance threshold (Figure 2). Considering the coordinates of significant ROC curves and further statistical analysis, we found an angle of 60.5 degrees as the best cut-off point for discrimination of high vs low in the interactive group of idiopathic dextroscoliosis scoliosis. In other words, in cases with idiopathic dextroscoliosis, the rate of SCT is significantly higher when the Cobb angle is above 60.5 degrees in comparison to the patients with lower Cobb angles. This cut-off value can be suggested as the best predictor of an underlying occult SCT in cases with idiopathic dextroscoliosis (Figure 2C; AUC = 0.68, P-value = 0.04, CI = 0.53-0.83, sensitivity = 85.7% and specificity = 43%). Accordingly, the 46.5 degrees Cobb angle is calculated as the cut-off value for congenital levoscoliosis although it did not reach the significance threshold due to low sample size (Figure 2E, P-value = 0.079, CI = 0.004-0.48, sensitivity = 57% and specificity = 90%).

**Figure 2:**
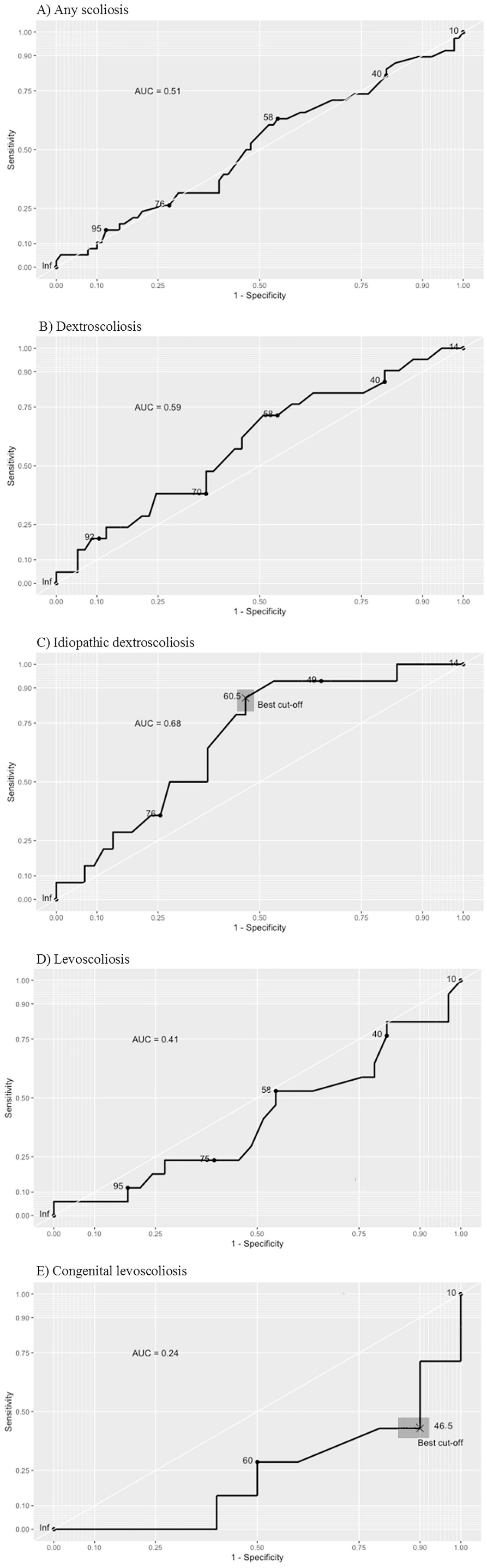
Operator Characteristic Curve (ROC) curve analysis of Cobb angle and spinal cord tethering in A) all scoliosis sample B) dextroscoliosis C) interactive subgroup of idiopathic dextroscoliosis D) levoscoliosis and E) interactive subgroup of congenital levoscoliosis

Interactive subcategories of scoliosis show various rates of underlying SCT. In our sample of cases, the lowest frequency of SCT was found among patients with low-angle (Cobb angle < 60.5°) idiopathic dextroscoliosis (8%). On the other hand, in cases with low-angle (Cobb angle < 46.5°) congenital levoscoliosis, the rate of SCT was highest (80%). As for the low-angle congenital levoscoliosis subgroup, we found that the rate of SCT is significantly higher, compared to any other idiopathic subgroups. (Figure 3).

**Figure 3:**
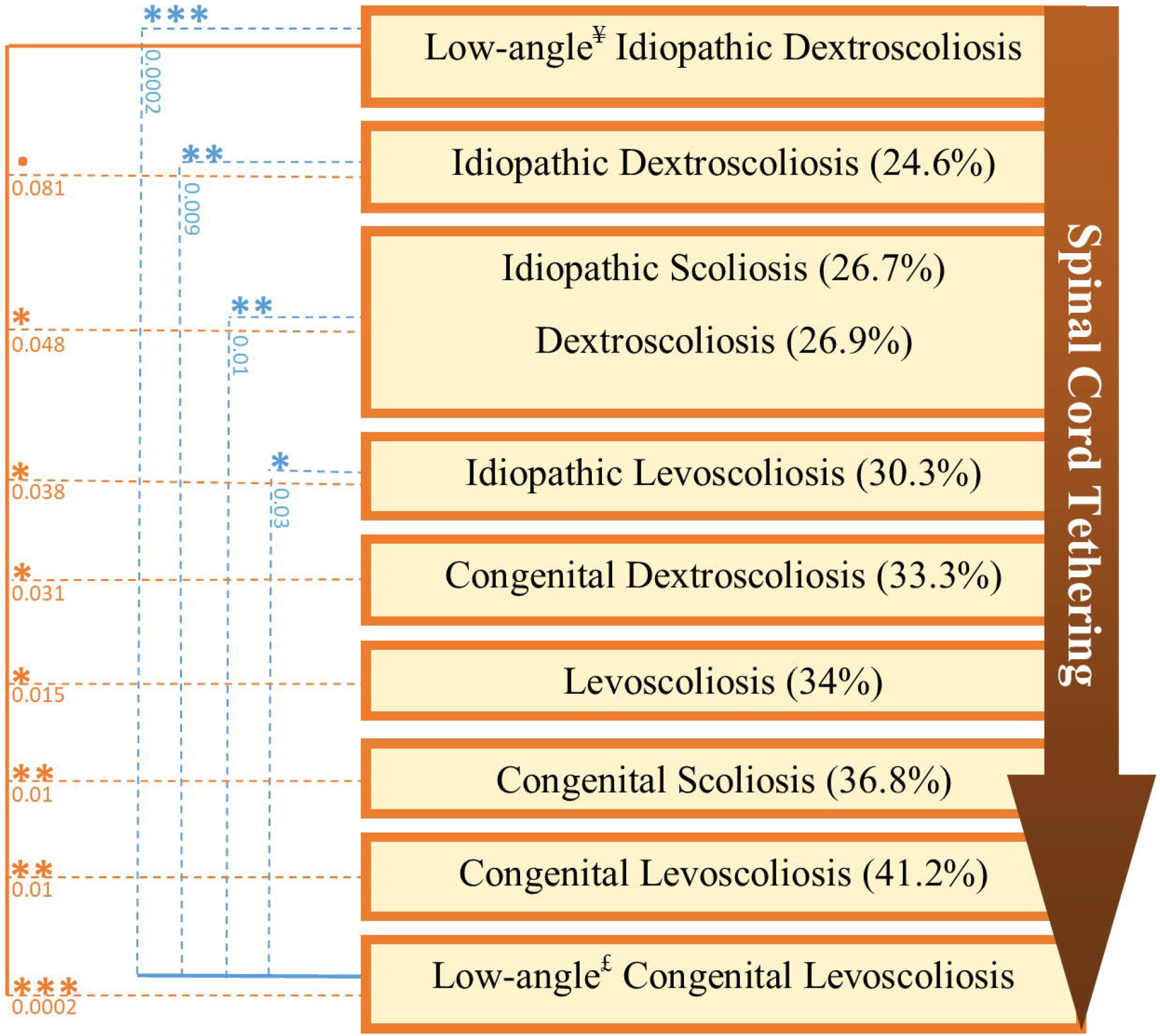
Flow diagram comparing the frequency of spinal cord tethering in some of the interactive subcategories of scoliosis. [£ = Cobb angle less than 46.5; ¥= Cobb angle less than 60.5]. We sorted the subcategories of scliosis based on the probability of an underlying spinal cord tethering. The asterisks show the level of statistical difference and underlying numbers represent the responsible P-value.

## Discussion

In this study, we evaluated the frequency of underlying SCT in different subcategories of scoliosis based on its characteristics. The main finding of this study is that higher Cobb angles in idiopathic dextroscoliosis cases increase the likelihood of the presence of an underlying cord tethering abnormality. On the other hand, it could also be concluded that with the presence of SCT in cases of idiopathic dextroscoliosis, a higher Cobb angle and more severe deformity can be expected. This relationship was somewhat reverse in congenital levoscoliosis, i.e., lower Cobb angles seem to have an association with SCT. No subgroup could be detected with no risk of spinal cord tethering.

### Interpretation of general findings

To check for the interpretability and generalizability of this model, we compared our sample’s clinical and demographic features with other reports. Among our participants, 39.1% had levoscoliosis and dextroscoliosis was found in 60.9%. This was similar to a previous report with larger sample size. .(10) Almost 70% of our cases had idiopathic scoliosis, which is comparable to 80% reported in another study. (4) Sixty-eight percent of participants were females. This is in accordance with previous studies, demonstrating that scoliosis and many other neurosurgical disorders are more commonly found in females or it might be related to access and knowledge toward healthcare utilization. (30-32) A M:F ratio of 1:1.5 in dextroscoliosis cases and 2.7:1 for levoscoliosis patients, is similar to other works,(4, 15, 21, 33, 34) marking that male patients’ spinal column is more likely to deviate to the left. Overall, it seems that this model can represent the general population of scoliosis patients. It should be noted that in infantile scoliosis, such a propensity in male cases has not been reported.(22)

Although SCT was more commonly found in levoscoliosis, no statistical difference was found regarding the frequency of SCT between levo- and dextroscoliosis spinal column configurations. These findings have been supported by previous works.(21-23) Similarly, in another study the rate of spinal cord anomalies was higher in levoscoliosis but the difference was not found to be significant. (23) The same is true for evaluating the effect of type of scoliosis on the frequency of spinal cord anomalies. Although these anomalies are more commonly found in congenital scoliosis, the difference is not significant.(3, 14, 23, 33) Considering the number of cases in our study and previous ones, it should be emphasized that the difference may become significant if the total sample size increases to enough number for each subgroup. Thus, further works with larger sample sizes and multicenter studies can be helpful in this regard.

### Tethered cord and structural features of scoliosis

Previous researchers have demonstrated that patients with underlying anomalies (including both tethering and non-tethering causes) do not have greater Cobb angles.(3, 14) Similarly, our results indicate that when the entire cases of scoliosis are considered, the rate of SCT does not significantly differ when the Cobb angle increases. This notion comes into interest when mentioning that this behavior is true for dextroidiopathic patients and completely reverses in levoscoliotics. Using the method described before, a cut-off angle of 60.5° in idiopathic dextroscoliosis is suggested as the optimal number to dichotomize idiopathic scoliosis patients with right-bending columns into low-angle and high-angle. It is emphasized that these cut-off angles are only proposed as practical values for prediction of SCT not for surgical planning of scoliosis and as a clue for pre-surgical awareness of tethering pathologies. A reverse pattern of the ROC curve in levoscoliosis patients suggests that an increase in Cobb angle not only is not a good predicting factor for the presence of SCT but also may play a protective role for it. A conviction with no known reason to this point.

Interactive subcategories introduced in this study may help the surgeon to better be prepared for pre-surgical planning and intra-operative alertness. The lowest frequency of SCT can be found among patients with low-angle (Cobb angle < 60.5°) idiopathic dextroscoliosis (8%). On the other hand, patients with low-angle (Cobb angle < 46.5°) congenital levoscoliosis, though rare, had the highest rate of SCT (80%). Within our subgroups, the most common configuration was high-angle idiopathic dextroscoliosis. Twenty-five percent of our entire cases were in this subgroup, having a 37.5% rate of SCT.

There are many authorities, who believe that pre-operative lumbosacral MRI is not necessary in every case of scoliosis. Especially in idiopathic cases with dextroscoliosis, the debate is present. Many institutions do not routinely perform pre-operative MRI and rely on certain criteria in determining whether MRI is indicated. But as it can be identified, even in the lowest risk subgroup of ours, the possibility of an underlying spinal cord tethering is not zero. On the other hand, for patients ath higher risk of tethering, such as a dextroscoliosis patient with high Cobb angle, a normal-appearing conus can be misleading in convential pre-operative MRI and some tetherings might be overlooked. In such cases additional modalities such as urodynamic study (18), diffusion weighted MRI (35), or prone MRI (27) can be utilized for exclusion of SCT.

The exact reason why the relationship between Cobb angle and the frequency of SCT is not identical in idiopathic dextroscoliosis and congenital levoscoliosis is obscure. It can be hypothesized that in some cases of idiopathic dextroscoliosis, SCT is the main cause for the anomaly, which can also produce more severe deformities; while in congenital forms, other genetic factors play the cardinal role in producing scoliosis. Possibly, the presence of SCT manifestations urges the parents to seek medical advice in earlier phases of deformity. Perhaps, in the congenital levoscoliosis group, this is the main reason why the rate of SCT is more in patients with lower Cobb angles. In other words, the positive relationship between Cobb angle and the risk of SCT in idiopathic dextroscoliosis may be due to the causational effect of SCT on the development of deformity in idiopathic cases, while in congenital levoscoliosis, genetic alterations may cause both SCT and deformity and the patients with symptomatic SCT tend to be visited earlier by the physician and at lower Cobb angles. We cannot certainly claim such causations through the results of our study and a firm conclusion cannot be drawn, but this can be a subject to consider for further investigations.

### Limitations

Several limitations of the study need to be considered: 1) referral nature of our clinic makes selection bias possible and the results should be used cautiously 2) the cross-sectional design of this study and its relatively low sample size are other possible shortcomings. Further studies with a larger sample size are needed to better evaluate the associations.

## Conclusion

Higher Cobb angles in idiopathic dextroscoliosis cases are statistically associated with a higher frequency of underlying SCT. In contrast, in congenital levoscoliosis patients, lower Cobb angles may highlight the higher rate for cord tethering. The lowest frequency of cord tethering can be found in patients with low-angle idiopathic dextroscoliosis, but even in this subgroup, the rate of SCT is not zero. A surgeon should be aware of tethering pathologies regardless of type or severity of scoliosis and MRI should be preserved as a routine test for every case of scoliosis before operation, regardless of Cobb angle, type of scoliosis, and convexity. On the other hand, we recommend strict caution and a high level of suspicion in the evaluation of the high-risk subgroups (e.g. low-angle congenital levoscoliosis group) since the risk of spinal cord tethering is dramatically high in these cases. In other words, a clean pre-operative conventioal MRI may not be enough to rule out tetherings of spinal cord in selected patients at high-risk of SCT and further work-ups such as urodynamic study, prone MRI, or unconventional sequences (e.g., diffusion weighted) might be better determinants of tethering. A matter that needs to be evaluated in future works.

## Data Availability

All data produced in the present study are available upon reasonable request to the authors

## Disclaimer statements

### Funding

No financial support has been allocated from any organization.

### Conflict of Interest

Authors declare no conflict of interest

### Availability of data and material

All the material and data used in this manuscript are available upon request to the corresponding author

### Statement of ethics

This study was approved by the Institutional Review Board of our hospital. Both written and oral explanations of the contents of this study were provided to the participants, and their consent was obtained. We followed the STROBE guideline for reporting observational studies.

## Notes

### Competing Interest Statement

The authors have declared no competing interest.

### Funding Statement

This study did not receive any funding

### Author Declarations

The ethics committee of Tehran University of Medical Sciences gave ethical approval for this work.

